# Menstrual hygiene management improvement in selected communities using nurturing care group approach

**DOI:** 10.1101/2024.10.01.24314753

**Authors:** Bismark Dwumfour-Asare, Eugene Appiah-Effah, James Ben Tidwell, Kwabena Biritwum Nyarko

**Affiliations:** Department of Environmental Health & Sanitation Education, Faculty of Environment and Health Education, Akenten Appiah-Menka University of Skills Training and Entrepreneurial Development, Asante Mampong Campus, Ghana; Regional Water and Environmental Sanitation Centre Kumasi, Department of Civil Engineering, College of Engineering, Kwame Nkrumah University of Science and Technology, Kumasi, Ghana; International Programs Group, World Vision US, Federal Way Washington, US.

**Keywords:** challenges, education, menstrual hygiene, rural settings, stigma, taboos, women

## Abstract

Improved menstrual hygiene management (MHM) is key to socioeconomic empowerment of women through improved well-being. Safe MHM is challenging in developing countries especially in rural and low-income settings. The study assessed the influence of World Vision’s piloted project of Nurturing Care Group approach on MHM in rural communities of Ghana. Four (4) communities each from Savelugu Municipal and Sekyere East District Assemblies were selected for a cross-sectional qualitative survey using 16 focus group discussions. Data was analysed using inductive mixed method with content analysis. Findings from 162 study participants showed satisfactory MHM awareness and practices among young and old females; MHM awareness and education sources included relatives, friends, NGOs, public and social institutions (e.g., health centres, churches, schools etc); intervention improved MHM understanding, behaviour and personal hygiene practices. Some MHM challenges that persisted included non-availability and prohibitive high cost of MHM materials, burden of managing menstrual pains, inadequate knowledge in handling menstrual intricacies, menstrual taboos, and stigma. Menstrual stigma was comparatively low among intervention communities than controls, but there was no difference regarding menstrual taboos across all communities largely due to religious and superstitious beliefs. Championing local production and use of reusable menstrual pads from fabrics was considered an innovation to ease sanitary pad cost burden and non-availability of menstrual pads. MHM was positively influenced through improved understanding of personal hygiene practices, and reduced stigmatization among others. Subsequent interventions would require adequate scope and resources to work with stakeholders to facilitate change processes with persistent MHM challenge like taboos.

## 1 Introduction

Menstrual hygiene management (MHM) is relevant to achieving the sustainable development goals (SDGs) linked to i) SDG 3 – good health and well-being, ii) SDG 4 – inclusiveness and quality education, iii) SDG 5 – gender equality and women’s empowerment, SDG 6 – clean water and sanitation, and SDG 8 – decent work and economic growth. These goals are key to in addressing the needs of women, girls, and especially those in vulnerable situations. People who menstruate face multiple challenges in managing their menstruation in homes, schools and workplaces (1). The challenges are compounded by factors like long distances from home to school, poverty, and lack of facilities (water, toilets and changing rooms) etc especially in rural areas (2–4). Others include lack of menstrual materials, menstrual stigma and taboos, period pains and other associated menstrual illness (distress, dysmenorrhoea, headache and excessive bleeding) (5, 6). Thus, dignity and well-being of menstruators in low-income settings are affected by the quality of menstrual hygiene management (7). Yet, menstruators face challenges due to little knowledge about menstruation and/or lack of information on safe menstrual hygiene practices (8).

Menstruators exist in all settings worldwide, yet menstruation is still shrouded in silence in many cultures. The stigma associated with menstruation has been universal whether in Asia, Africa, Australia and America (9). Some cultures, like Nepal and the Pacific, still implore women to stay in secluded locations like menstrual huts during their period (10, 11). Some restrictions, such as avoiding household chores are perceived as desirable by women (12). For instance, among the Akans in Ghana, women are faced with multiple restrictions against certain engagements such as visiting shrines, attending palace hearings, and cooking for husbands (and/ or male household members) when menstruating, emphasizing the existence of social seclusion of women (13).

Urgent attention is therefore needed on the challenges of MHM such as articulated earlier including stigma and taboos (and the fear thereof), poor hygiene practices, lack of antidote for menstrual pains and other illnesses (14–16). MHM constraints create menstrual poverty including other restrictions like inadequate education and training (17), and teasing adolescent girls with period stains and/ or odour (18). Menstrual poverty also limits socio-economic empowerment in low and middle-income countries (19). Thus, for healthy and productive lifestyle, and dignified lives, addressing practical concerns is key (20).

Recent campaigns for improved MHM emphasise positive period and female empowerment (21). Efforts to change the narrative about MHM are pushing interventions to break menstruation stigma and taboos by providing relevant knowledge, skills and resources (22). Interventions provide opportunities to women, girls and communities to speak about menarche and period myths which are intrinsically linked to local menstrual taboos (12, 17).

World Vision piloted the Nurturing Care Group (NCG) approach in Ghana from June 2019 to January 2021 in two (2) programme areas – Savelugu Municipal Assembly and Sekyere East District Assembly. The NCG approach has been described in detail in a forthcoming paper on animal faeces management (AFM) (23), a core of the two project components in the NCG intervention. Moreover, the model was based behaviour change approach which empowered volunteer group leaders to create a multiplying effect to reach households through neighbour-to-neighbour contacts, home visits and group meetings (24, 25). The NCG pilot project was designed to focus mainly on improving MHM and AFM among others in selected communities. The target was addressing MHM and AFM challenges in home and community management through collective action for health and development, safe water, sanitation and hygiene etc. (24). MHM and AFM are usually neglected in environmental sanitation and/ or WASH interventions in communities.

The qualitative evaluation study of the NCG pilot project is split into two separate papers because of the complexity of concept presentations making it impossible for a single clear and digestible integrative article, and contrasting scope and themes of project components with distinct attributes (26). This paper on one part presents the qualitive assessment of the MHM component of the NCG pilot project while the other forthcoming paper, Dwumfour-Asare et al. (23), shares the findings on the AFM. This study reports on the effectiveness of the MHM component in terms of knowledge transmission, behaviour change practices, and changing social norms (breaking stigma and taboos) and yet prevailing barriers in the study communities.

## 2. Methodology

### 2.1 Description of the Study Area

The study was undertaken in the two districts - Savelugu Municipal and Sekyere East District Assemblies in Ghana. While Savelugu Assembly is dominated by the Mole-Dagbani (88.4%) ethnic groups, and Moslems (95%), the Akans are the majority tribe (85%) with most worshippers (76%) being in Sekyere East District (27, 28). More information about the study areas including some sociodemographic data is shared in details in the paper on animal faeces management (AFM) component of the NCG pilot project (23).

### 2.2 Study Approach and Data Collection

A cross-sectional qualitative survey was carried out in communities with and without the NCGA project intervention. The data collection campaign took place between 22^nd^ November 2021 and 11^th^ December 2021 in 8 communities: 4 each from Savelugu Municipal and Sekyere East District Assemblies. The survey involved 16 focus group discussions (FGDs) with two main women groups (those above 25 years old, and those below 25 years i.e., 15 – 25 years) in each selected community. In each district, two intervention communities (1 successful and 1 less successful based on animal faecal management component of the intervention) and two control (non-intervention) communities were randomly selected. Females who were in the community at the time of the NCG intervention were recruited as study participants based on their availability at the time of the survey by random hand-picking by community leaders and/ or care group leaders.

The FGD sessions were facilitated using FGD guide with questions based on the key themes: background of the NCG program, participants’ sources and kind of information on MHM before intervention, specific knowledge acquired/lessons recall, lesson spill over from intervention communities to non-intervention areas, and existing barriers to MHM. The FGDs were held using the two local languages of participants like the forthcoming AFM paper (23). Two qualified translators were recruited with the support of the Savelugu World Vision Area Program Office. The FGD sessions took between 1 and 1.25 hours to complete with audio and field notes recorded.

### 2.3 Data Management and Analysis

First, data from the FGDs were transcribed and organized using Microsoft Word similar to the AFM paper (23) adapting the three-stage approach to minimize errors – using audio recording and field notes together by careful listening before transcriptions (29). All transcribed data were finally transferred to Microsoft Excel database for data analyses following key terms for trends and prevalence, and inductive mixed method with content analysis, including careful identification of emerging topical themes and supporting synthesis with verbatim quotes (30, 31).

### 2.4 Ethical Considerations

Ethical clearance certificate was secured from the Committee on Human Research and Publications Ethics (CHRPE) of the Kwame Nkrumah University of Science Technology was sought with reference number Ref: CHRPE/AP/418/21 before field work was executed. All approved protocols were followed including informed consent from participants after reading and explaining information about the study to them. The verbal informed consent from participants was witnessed by the leaders who led participants’ recruitment and the consent was documented as part of the field recordings (audio and field notes).

### 2.5 Limitations of the study

The key limitations are that – the findings are limited to the few selected communities in the two pilot district assemblies. The study is focused on tracking the immediate effect or influence of the project intervention to understand some relevant lessons from participants’ or beneficiaries’ perspective. The study is not an impact evaluation of the NCG pilot project but an assessment to understand any immediate influence felt in communities and what did not work well to inform future interventions.

## 3. Results

### 3.1 FGD Participants

The study participants communities and participants were mainly subsistence farmers. Table 1 presents the two (2) main FGD categories – women under 25 years, and those above 25 years from four (4) intervention and four (4) control communities. The group sizes ranged from 6 to 15 participants, with an average of 10 members per focus group. The total participants involved in the study were 162 women (young and old).

**Table 1:** Number of participants for the FGDs in the study.

| District | Community | Type of community | Participants of Focus Groups |  |
| --- | --- | --- | --- | --- |
|  |  |  | <25 years | >25 years |
| Savelugu | Gbumbum | Intervention | 9 | 11 |
|  | Zieng | Intervention | 6 | 8 |
|  | Naprisi | Control | 8 | 9 |
|  | Kukobila | Control | 7 | 7 |
| Sekyere East | Asukokor | Intervention | 14 | 14 |
|  | Feyiase | Intervention | 7 | 15 |
|  | Nsutam | Control | 10 | 12 |
|  | Tetekaso | Control | 11 | 14 |
| **Total** |  |  | **72** | **90** |

Table 2 presents the results of thematic analyses of responses on sources of menstrual awareness and education; challenges with menstrual hygiene management; and existing menstrual stigma, taboos and myths.

**Table 2:**
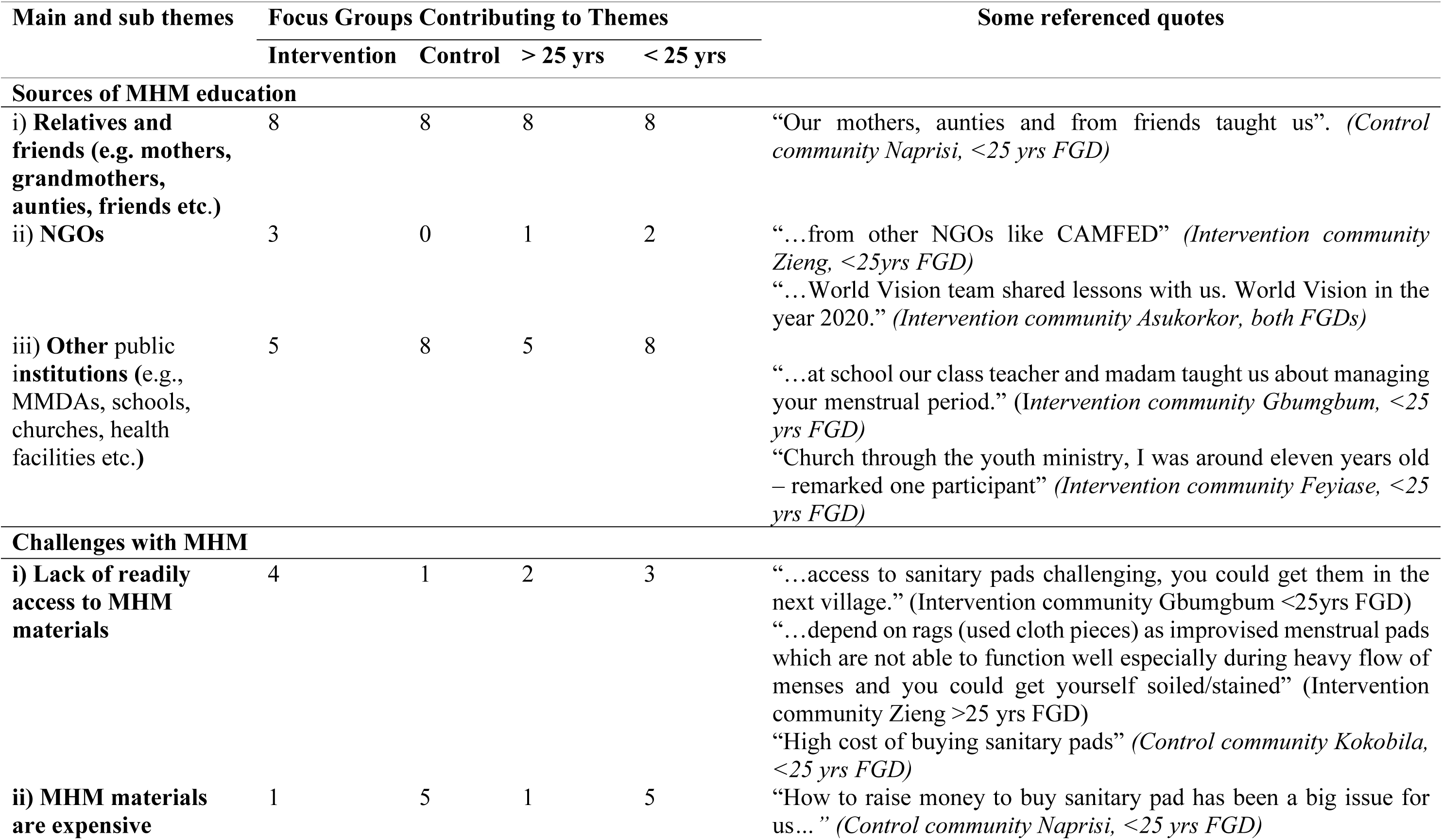
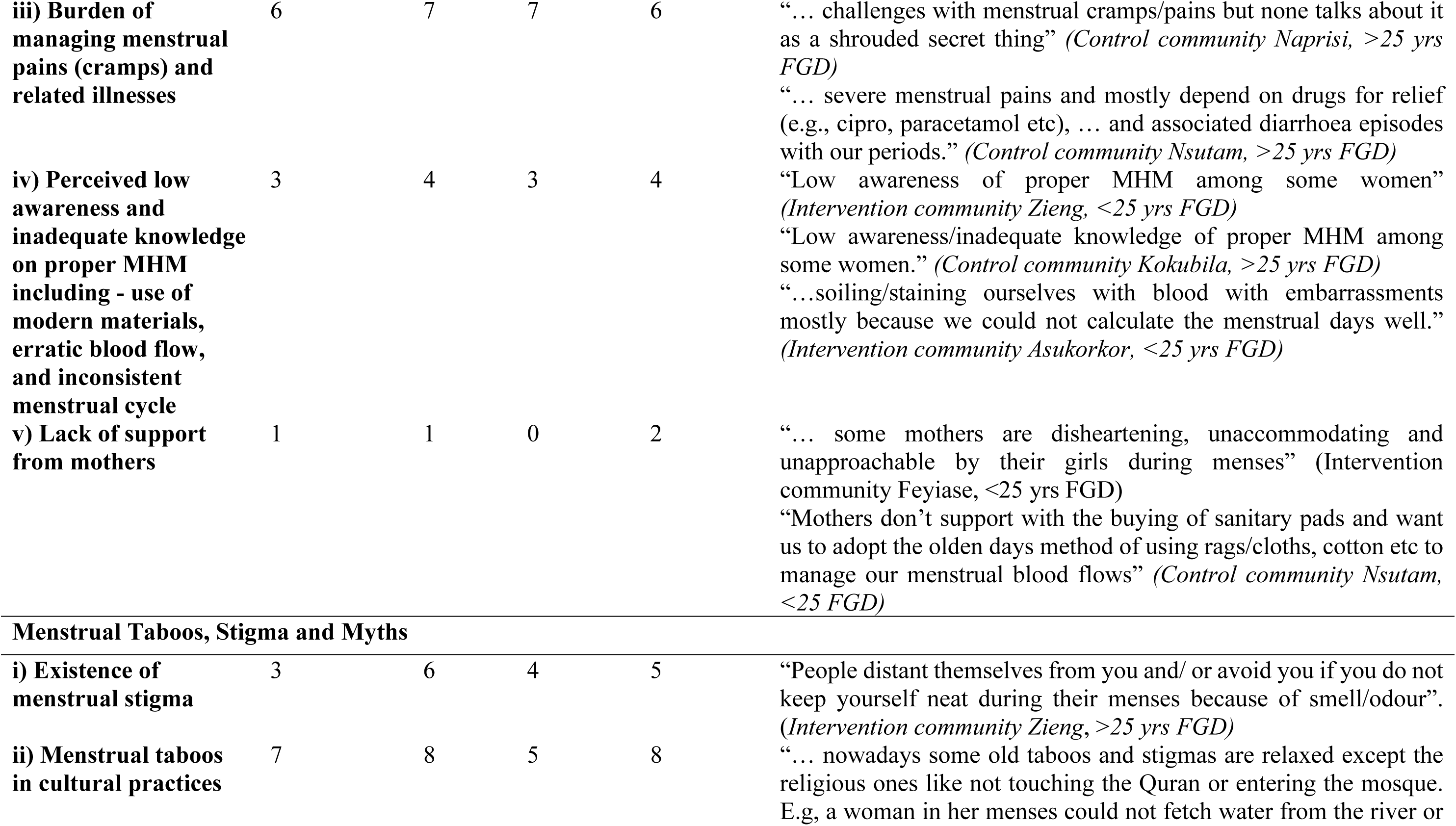
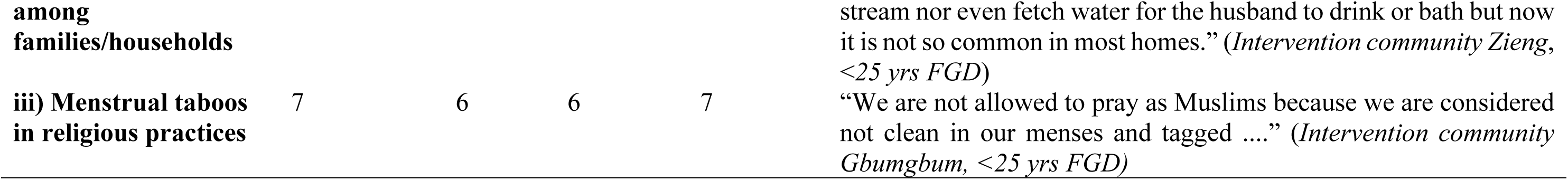
Key and sub themes analyses from the focus group discussions.

Participants largely received awareness and education on MHM especially for the first time from relatives and/ or friends, and subsequently from NGOs, and other public institutions. The challenges with MHM were mostly period poverty – attributed to lack of access to MHM materials, and inability to afford materials, followed by the burden of managing menstrual pains and related illnesses. Menstrual stigma and taboos existed in the communities, and these were embedded directly or indirectly in religio-cultural beliefs and practices including myths. The results showed mixed trends in terms of differences between intervention and control communities and across age groups.

### Sources of menstrual education

Study participants cited that menstrual education were mostly spearheaded by females. The education sources included social institutions like families and friends, community neighbours, schools, health centres, churches etc (Table 2). Within the family, menstrual education was done by grandmothers, mothers, older female siblings, and other close relatives who had experienced menstruation themselves. Information from health centres in communities was mentioned to be useful especially on managing menstrual pains and maintaining personal hygiene. The findings were mostly collective responses and/ or similar experiences shared by individuals which were confirmed among group members.

Some participants disclosed the following:

> *“My mother discussed it with me” “… my senior sister discussed it with me” (Participant, <25 yrs FGD, Gbumbum)*

For some participants, MHM was learned by paying attention to the practices of older people as cited:

> *“I learnt from siblings especially by observing my older sisters’ practices” (Participant, <25 yrs FGD, Feyiase).*

> *“I learnt through attentive listening from one lady who mostly sent me to buy sanitary pads and cotton wool” (Participant, <25 yrs FGD, Nsutam).*

Interestingly, two young participants did not get the opportunity to learn from anyone especially an adult female relative but through their own personal experiences. One had lived with her father alone and recounted that her father was never informed about her menarche because she felt shy.

> *“I was shy to tell my father when living alone with him as single parent” (Participant, <25 yrs FGD, Feyiase).*

> *“I never had the opportunity to be taught by anyone close to me. I learnt it on my own in a hard way – I had to do something the first time and afterwards by personal experience.” (Participant, <25 yrs FGD, Asukokor).*

Meanwhile, some responses indicated that transfer of MHM knowledge from adults to the young people was generally low and some mothers did not consciously share any menstrual information with their female wards.

> *“Some of us have discussed MHM with our children but others have not because they believe or assume their children already know it, and they hide it from them.” (Intervention community Asukokor, >25 yrs FGD)*

> *“Attitude from some mothers is disheartening; they are unaccommodating and unapproachable by their daughters” (Intervention community Feyiase, <25 yrs FGD).*

> *“Mothers don’t support with the buying of sanitary pads and rather want us to adopt the olden days method of using rags/cloths, cotton wool etc. to manage our menstrual blood flows” (Control community Nsutam, <25 yrs FGD)*

> *“We need to be supportive to our children (young girls) when in their menses” (Control community Tetekaso >25yrs FGD)*

After the intervention, some participants indicated improvement in mothers relating well with their young girls, however some young participants did find any significant paradigm shift in communicating and/ or sharing menstrual management information from adult women. Some participants of the young FGD had these to say:

> *“We see our mothers holding books and making rounds, but they never reach us with any education or information. …. we see that the mothers only take their books when they hear that World Vision group is coming around.” (Intervention community Feyiase <25 yrs FGD)*

### Menstrual hygiene management challenges

Study participants expressed several views across groups on their challenges during menstrual periods. However, the challenges raised were similar (Table 2), which included low awareness and access to modern MHM materials like sanitary pads, menstrual pains/cramps, lack of self-confidence during menses etc. MHM stigma and taboos existed as challenges but have been considered in the next section in details. Meanwhile, the incidence of the challenges were relatively pronounced in the control communities, and among the younger participants.

For control communities, participants were emphatic with the following challenges:

- low awareness and inadequate knowledge of proper MHM,
- erratic and inconsistent menstrual cycles,
- menstruation related health conditions like menstrual cramps/pains, diarrhoea, skin rashes etc meanwhile the related health issues are shrouded in secrecy and not discussed/talked about,
- high cost of MHM materials like sanitary pads,
- overuse of pain killers (e.g., cipro, paracetamol etc.),
- improvisation of toilet tissue paper instead; and
- lack of support from parents especially mothers in providing MHM materials, etc.

> *“The challenge is how to manage menstrual cramps/pains but unfortunately people don’t talk about it because it’s considered a secret thing. … challenges with menstrual cramps/pains but none talks about it as a shrouded secret thing” (Control community Naprisi FGDs)*

> *“…my experience is frequent visits to the toilet – diarrhoea episodes when in I am in my menses.” (Participant, Control community Nsutam, >25 yrs FGD).*

> *“… severe menstrual pains and mostly depend on drugs for relief (e.g., cipro, paracetamol etc), … and associated diarrhoea episodes with our periods.” (Control community Nsutam, >25 yrs FGD)*

> *“Mothers don’t support with the buying of sanitary pads and want us to adopt the olden days method of using rags/cloths, cotton etc to manage our menstrual blood flows” (Participant, control community Nsutam, <25 FGD)*

For intervention communities, the following were the MHM challenges:

- Low awareness and access to modern MHM practices including materials like sanitary pads although some are uncomfortable to use,
- Menstrual pains/cramps and headaches including bodily weaknesses and associated illnesses,
- Lack of self-confidence during menses,
- Erratic blood flow and inability to predict menstrual cycle which may come with public embarrassment when stained with blood, and
- Inadequate parental support to young females on MHM, etc.

> *“Getting access to a sanitary pad is challenging. Pad is not sold in our village, so we have to travel to a nearby town to get one.” (Intervention community Gbumgbum, <25yrs FGD)*

> *“We largely depend on rags (used cloth pieces) as improvised menstrual pads which were not able to function well especially during heavy flow menses and you could get yourself soiled/stained.” (Intervention community Zieng, >25 yrs FGD)*

> *“…menstrual pains and headaches are common with young girls in this community during menstrual periods.” (Intervention community Feyiase, <25 yrs FGD).*

> *“…soiling/staining ourselves with blood with embarrassments mostly because we could not calculate the menstrual days well.” (Intervention community Asukorkor, <25 yrs FGD).*

The older group were much particular about menstrual pains challenges and their inability to manage the situation nor get any support from anywhere. Frustrations were worsened by stress and depressions associated with menstrual cramps crises combined with over-burdened domestic chores. These negatively affected self-esteem and confidence to meet public expectation.

> *“…how to behave comfortably in public during menstruation.” (Participant, >25 yrs FGD Gbumbum).*

> *“… any challenges are hardly talked about, if any at all, except menstrual pains.” (Intervention community Asukokor, >25 yrs FGD).*

> *“Sometimes the menstrual period changes, and you may miss the timing, which could lead to emergency blood stains in public with embarrassment. Mostly this happens when we are not able to do the calculations well.” (Intervention community Asukokor, < 25 yrs FGD).*

Aside these challenges, young participants in the intervention communities expressed their dissatisfaction with the attitudes of their immediate family members towards them including mothers. Also, some mothers had pushed their daughters to use improvised MHM materials like rags and tissue papers other than modern sanitary pads.

> *“…the attitude from some mothers is not good, they are unaccommodating and unapproachable by their daughters.” (Intervention community Feyiase, <25 yrs FGD)*

### MHM Taboos and Stigma

Menstruation associated taboos and stigma were driven by religious beliefs, traditions, lapses in personal hygiene practices of victims, and to some extent exposure to modernization. Menstrual taboos and stigma were similar across communities irrespective of the demographic differences (north versus south, and young versus old). Thus, in Savelugu (northern Ghana) with dominant Muslim communities, restricting women in menses from religious activities like joining congregational prayers and/ or attending worship at the mosque, contacts with and/ or serving males (e.g. husbands, fathers etc) in homes were no different from similar practices in the Sekyere East (southern Ghana) where same exclusion was reported with spiritual churches, shrines and fetish priests, contacting and/ or serving males/husbands, visiting some rivers/streams etc.

FGD participants from the north shared their views as follows:

> *“We women are not allowed to pray as Muslims at the mosque because we are considered unclean during our menstrual periods” (Intervention community Gbumbum, >25 yrs FGD).*

> *“… women in menses could not touch the Holy Quran and could not join prayers at the mosque too because they are considered unclean at that time.” (Intervention community Zieng, <25 yrs FGD)*

> *“… women can’t have sexual intercourse or any intimacy with partners during our periods.” (Intervention community Gbumbum, >25 yrs FGD).*

> *“We are not allowed to pray and do not have sexual intimacy with our partners.” (Intervention community Kukobila, >25 yrs FGD).*

In the southern communities of Sekyere East, some experiences were as follows:

> *“… women cannot go to a fetish priests’ houses or shrines in their menses, even very daring to pass by such houses/shrines or greeting them.” (Intervention community Feyiase, <25 yrs FGD)*

> *“Some churches don’t allow you to come to church, especially the spiritual churches/movements.” (Intervention community Asukokor, <25 yrs FGD)*

> *“… we do not cook nor hold/touch our father’s food.” (Intervention community Asukokor, <25 yrs FGD).*

> *“You dare not cook for your father nor touch his drinking cup.” (Control community Tetekaso, <25 yrs FGD).*

The older participants expressed worry that menstruation taboos affected female lifestyle including domestic chores, for instance, inability to fetch water, cook and serve food to their husbands and male household members, visit certain farmlands, nor commuting over rivers/streams for socioeconomic ventures etc. Some menstrual taboos could be extensive barriers including restrictions on commuting the only available route of riverine transport.

> *“… we are not allowed to fetch water from the dam.” (Control community Kukobila, <25 yrs FGD).*

> *“…you don’t cross certain rivers nor attempt to access or fetch water from such rivers in the community.” (Intervention community Feyiase, <25 yrs FGD).*

> *“…you don’t have to commute across certain rivers in the community, including going to farms and markets.” (Intervention community Asukokor, >25 yrs FGD).*

Some menstrual taboos were observed because of the belief in the dire repercussions on women who flout them. In both Savelugu and Sekyere East, some participants shared their personal experience to consolidate the fear of menstrual taboos, citing consequences such as prolonged and/ or heavy flow menses.

> *“… I bled continuously for almost two weeks after I crossed that river down the community to my farm, although I didn’t know about the taboo” (Participant, Intervention community Asukokor, >25 yrs FGD).*

> *“The taboo is real, and some of us have suffered the consequence before, that if you defiled it you experience protracted menstrual periods for several days to weeks.” (Control community Kukobila, >25 yrs FGD).*

On menstrual stigma, some participants cited that teasing by boys in schools was common although culprits were liable to punishments when reported to school authorities to prevent such stigmatization.

> *“When a girl stains herself in school, they may call her names, especially by the boys, saying Osmo! Osmo! Stigmatization may happen only at school, especially when you soil yourself with blood stain – this normally comes from the boys, although they could be punished when it is reported to school authorities.” (Control community Nsutam, <25 yrs FGD)*

Although, adults were not subjected to menstrual stigmatization, they recalled experience including schooldays, confirming the long-persisted ordeal in communities.

> *“Stigmatization was common at school in those days when you got your dress stained.” (Control community Nsutam, >25 yrs FGD)*

> *“People distant themselves from you ….during their menses because of smell/odour.”*

(Intervention community Zieng, >25 yrs FGD)

### Behavioural changes to MHM Promotion

For control communities, information and education on MHM came from sources other than the NCG intervention and focused on use of modern sanitary pads, improved personal hygiene practices, and supporting others especially young females.

The key messages from Savelugu control communities were:

✓ Using modern sanitary pads, appropriate pad usage practices, and improved personal hygiene like regular bathing – at least twice a day.
✓ Stop taking family planning pills during menstrual periods for effective monitoring of menstrual cycle.

> *“Previously we could use one sanitary pad the whole day, but now we change pads at the right time. You can get infections and sores from using one sanitary pad for a long time.” (Control community Naprisi, <25 yrs FGD)*

> *“… we have not changed anything much except using modern pads for improved personal hygiene.” (Control community Kukobila, >25 yrs FGD)*

Likewise, participants from Sekyere East control communities shared the following:

✓ Preparing in advance towards menstrual episodes for safe MHM
✓ Foreknowledge that alternative MHM materials like clean fabric other than sanitary pads could be used especially when properly washed, dried and pressed before reuse.
✓ Supporting young girls on MHM through education

> *“For us who improvise the use of cloth as pads, we wash the materials and iron it neatly before reuse and it has helped a lot especially when you have no money to buy sanitary pads.” (Control community Nsutam, >25 yrs FGD)*

For the intervention communities, the list of behaviour change practices was relatively exhaustive likely due to additional understanding from the pilot project sensitization. However, the improved behaviour changes did not include taboos but practices that could break or avert stigmatization linked to personal behaviour and hygiene practices.

Most participants were of the view that menstruation taboos existed but were not as stringent as in earlier generations in some places likely due to modernization and some religious beliefs contradicting traditional and cultural beliefs and practices.

Key improved behaviour change identified from the Savelugu intervention communities were:

> ✓ Able to use sanitary pads now and episode of menstruations may go unnoticed by any third party. There has been a drastic changed from using old cloth (rags) as sanitary pads to a reusable fabric or disposable sanitary pads with improved hygienic practices such as regular changes and bathing.

> ✓ Keeping oneself away from sexual intercourse to avoid unwanted (teenage) pregnancy.

> ✓ Improved personal hygiene practices including washing of cloths, underwear, reusable pads etc and drying in the sunlight.

> ✓ Inferiority complex that menstruating women were not clean enough to mingle with others are dispelled to the barest minimum. With improved hygiene practices a menstruating woman can have her normal life like any other person.

> ✓ Improvement in making advanced preparation towards menses period including educating other younger women on MHM.

Key improved behaviour change identified from Sekyere intervention communities were:

> ✓ Disposal of used sanitary pads is no more a nuisance, improved management of used MHM materials. Keeping used menstrual pads safe (by burying, burning and/ or dropping into toilet) from recent ritualists who scout for them for “ritual or blood money”.

> ✓ Educating and monitoring our children especially the girl child to provide the needed support on MHM. Some of us now relate well with our young girls share their concerns and support them during the menstrual periods.

> ✓ Schoolgirls are now able to manage themselves well without soiling themselves during menstrual periods.

> *“Now most women wrap used sanitary pads in polythene bags and dispose into toilet or burn them. This practice has been helpful because you hardly see used pads in the streets of the community especially where you could find dogs littering/dragging/running them around.” (Intervention community Asukorkor, >25 yrs FGD)*

> *“We still hear of taboos but are not as religious as they used to be some years back, especially in the olden days.” (Intervention community Feyiase, >25 yrs FGD)*

On the taboos, some were becoming unpopular. For instance, of not crossing certain rivers in the community during menstruating has become unpopular in recent times. The change were attributed to modernism influencing some beliefs regarding menstrual hygiene, and the observation did not appear much different between the northern and southern communities.

One of the groups in Savelugu was quoted:

> *“… nowadays some old taboos and stigmas are relaxed except the religious ones like not touching the Quran or not entering the mosque. For instance, a woman in her menses could not fetch water from the river or stream and even fetch water for her husband to drink or bath, but now it is not so in most homes.” (Intervention community Zieng, <25 yrs FGD).*

In the Sekyere East communities, menstrual taboos were said to be merely ceremonial - some people take them lightly in recent times including worshippers of strict church sects because the modern lifestyle of using sanitary pads could conceal any hint menstruation.

> *“… many people menstruate and go to church without the leaders being aware because of the use of modern sanitary pads.” (Intervention community Asukorkor, >25 yrs FGD).*

> *“We think some taboos are watered down by modernization, especially because of improved and safe MHM practices of modern life and educational promotion.” (Intervention community Feyiase, >25 yrs FGD).*

> *“….some church beliefs and modernisation have relaxed some menstrual taboos and stigma.*

> *…women in their menses could not go to farm to defile the land or soil but nowadays it is not like that.” (Control community Naprisi, >25 yrs FGD).*

In addition to awareness promotion from the intervention, some other factors contributed to the improved MHM practices among intervention communities such as provision of school toilet facilities with changing rooms by the intervention, improved knowledge of MHM among schoolgirls, supply of MHM materials to schools by NGOs etc.

Savelugu communities shared experiences from intervention and other sources:

✓ Improvement in the usage of sanitary pads including changing regularly due to MHM education.
✓ Schools have made provisions for changing rooms for girls. E.g. improvising a room in the school computer lab as changing room where possible.
✓ In homes, bathrooms could now be used as changing rooms during menstruation without stigmatization.
✓ MHM materials like sanitary pads were made available to schoolgirls by some NGOs.
✓ Schoolgirls learn MHM from their teachers.
✓ Practiced safe disposal of wastewater from washed menstrual pads (fabrics) unlike hitherto open disposal.

Sekyere East communities shared experiences from intervention and other sources:

✓ Improvement in the toilet facilities in schools where girls could access changing rooms.
✓ Schoolgirls showed improved knowledge on proper MHM practices and hardly soiled themselves with menstrual blood.
✓ Some latrines with urinals and changing rooms have been provided at schools by World Vision to support children especially the schoolgirls.
✓ Improvement in mother-daughter relationship especially to provide support on proper MHM.

Some of the quotations from FGDs are cited as follows:

> *“Toilet and urinals in schools were provided by World Vision to ensure privacy especially in MHM to support schoolgirls ….” (Intervention community Feyiase, <25yrs & >25yrs FGDs)*

> *“We have changed from using old cloth as sanitary pad to using reusable and/ or disposable sanitary pads. We did not know how to use the pads properly …..we were bathing only in the evenings but now we bath 3 times daily during menstruation.” (Intervention community Gbumbum, <25 yrs FGD).*

> *“Schoolgirls are now able to manage themselves well without soiling themselves during menstrual periods. Improved awareness, knowledge, and behaviour in that regard has been helpful.” (Intervention community Asukokor, <25 yrs FGD).*

> *“Previously wastewater from washed pads(cloths) was disposed of in the open, but now it is disposed of in a pit. Clothes, underwear, and reusable pads were washed and dried in our rooms, but now we have been educated to dry them in the sun.” (Intervention community Gbumbum, >25 yrs FGD)*

> *“We support by educating and monitoring our young girls on MHM. Some of us now have cordial relationships with them to share and support their menstrual issues together.” (Intervention community Feyiase, >25yrs FGD).*

## 4 Discussions

The sources of education and/ or awareness about safe menstrual hygiene management for study participants were mainly from relatives and friends across all groups – young and old, intervention and control communities. This is consistent with other studies indicating the significant role mothers and close relatives (e.g., older sisters and grandmothers) play as primary source of information on menstruation (17, 32), although such information sources may be inadequate (33). Moreover, this was complemented by support from public institutions like schools, churches, health facilities etc and NGOs (except for control communities). Control communities could not have received adequate MHM education from NGOs because MHM interventions at the community level are usually neglected even in WASH projects (34), unlike in schools. The contributions from NGOs, public and social institutions including local government authorities are relevant as recounted in other studies (35, 36). In Ghana, school authorities educate pupils on MHM under the supervision of School Health Education Program (SHEP), and communities are supported by health centres under Ministry of Health (37, 38).

The main challenges with MHM in the study communities are similar to those known in literature – lack of access and high cost of MHM materials (e.g., disposal sanitary pads), burden of menstrual pains (cramps), low awareness and inadequate knowledge on MHM among local women, and the feeling from young women that mothers do not provide adequate support MHM (39–41). The contrast is in the low incidence of the first two reported challenges among intervention than control communities. This could be attributed partly to better exposure to the knowledge of using modern sanitary materials from MHM intervention and other factors like inability to afford sanitary pads especially among the young women, lack of readily access to service etc. (36). The intervention communities were exposed to improved ways of managing menstrual periods including empowering them with skills to sew reusable pads from fabrics, yet, the preference for modern disposable pads was high likely due to repulsion to the burden of washing and reuse (42). Lack of affordability especially among young women undermines MHM support from parents or guardians in communities which could be linked to menstrual period poverty – where access to MHM materials becomes an unmet need (36, 43). The older women have become used to reusable menstrual pads (fabrics) and therefore are less worried about the cost of sanitary pads. Clearly several challenges exist among menstruating women yet limited platforms exist for them to voice out, even inadequate avenues at the household and community levels (33, 44).

Menstrual stigma marginally exists in communities, with about twice prevalence rate among control than intervention communities, yet no clear difference exists between the young and older women groups. Probably every woman might have had first-hand and/ or a colleague’s bitter experience including deep-seated internalised experience and perception (33). The NCG approach like other targeted behaviour change interventions could have partly contributed to the minimized menstrual stigma in the intervention communities (35). However, menstrual taboos are strongly felt in cultural (traditional) and religious beliefs and practices across all communities and not any significant difference exist between the intervention and control communities. Almost every discussant irrespective of the district (geographical location), community, religion and culture accepted that menstrual taboos strongly persist in their communities especially restrictions related to religio-cultural beliefs and practices (45). The intervention could barely bring any improvement in this regard, likely because religio-cultural practices are complex, mostly sensitive, and may require more time and integrated approaches including rigorous active engagement with stakeholders (key leaders – community, religious groups, etc.) (35, 46). The expected approach to this complex issue is working with and not directly against/opposing existing socio-cultural beliefs and practices including menstruation-related myths and denial, and religious restrictions (46) across major belief systems like Christianity, Islam and Traditional Beliefs (45).

Existing menstrual restrictions (because of taboos) plus other factors affected household practices such as exemption of menstruators from helping with house chores e.g., cooking, fetching water, serving males etc; restricted sexual intimacy, and even making appearances at social gatherings. The sentiments about these issues are more pronounced in Savelugu (north – Muslims and traditionalist) than Sekyere (south – Christians and traditionalist) communities likely because of differences in religio-cultural settings. This is consistent with a similar study conducted in a neighbouring district with similar demographics in the north (17), considered as characteristics among predominant Islamic communities in the Sub-Saharan region (16). Sekyere East communities have some menstrual restrictions and taboos relaxed due to modernization and increased in church beliefs. Some participants see menstrual taboos as outmoded practices and yet believe in the “spiritual” repercussions with the ordeal individuals’ go through like protracted menstrual episode even if one ignorantly flouted certain mystified menstrual taboos. For the traditionalist, the belief of mystical ability of menstrual blood to deactivate charms and spiritual powers exist and such taboos must continue (37).

In these communities, the fear of taboos may continue to exist without scientific proof of the associated consequences although such occurrences are possible (47). Yet, interventions could take opportunities in the decreasing significance of menstrual myths with associated taboos and stigma due to improvement in knowledge and modernization, and this is consistent with other taboos (48). For instance, restrictions on sexual intercourse may come with benefits and could contextualised with scientific knowledge to disabuse the traditional dogma. Scientifically, such restrictions could be averting the spread sexually transmitted diseases that are implicated in unprotected sex during menses time (49). Communities should be opened to discussions on the consequences of menstrual taboos which often lead to internalised secrecy, shame, decreased mobility, inequity, and negative impact on confidence and self-efficacy of women (36, 50).

Episodes of menstrual cramps are common challenges all women groups face and almost everyone manages the condition by self-medication through the use pain relief drugs like paracetamol, ciprofloxacin etc. similar to other studies (51). Also, some participants especially from control communities claim using antibiotics to numb menstrual pains and avert infections. Minority of women also rely on traditional herbal concoctions as painkillers and/ or treatment. Clearly, menstruators could be abusing drugs out of misinformation and self-medication, a common practice recorded in literature (14), and this could be contributing to the high prevalence of self-medication among Ghanaians (52).

MHM experience and lessons learned are pronounced among intervention than control communities as expected due to extra information, resource materials and support from the project implementation other than the common awareness and promotion activities in communities. The common MHM awareness are typically focused on personal hygiene practices, safe use of sanitary pads, pre-preparations towards menstrual cycles, etc. (22). NCG communities’ improved exposure could added rich experience narrative such as: better MHM hygiene practices (e.g., bathing at least thrice daily, regular changing of pads, etc.), producing reuseable sanitary pads from fabrics, improvement in self-confidence, safe disposal of wastewater and used MHM materials, improvement in supporting younger women (daughters and sisters), paying much attention to MHM issues etc. Thus, interventions that empower women through improved knowledge and understanding are capable of influencing positive attitude and behaviour change practices (53), including innovation, self-confidence and efficacy, reducing anxiety and dispelling negative psychosocial attributes (22).

### Conclusions

There is a satisfactory level of menstrual hygiene management awareness and practices among females (young and old) in the study areas. Women in communities receive menstrual hygiene management awareness and education from varied sources including relatives and friends, NGOs, public and social institutions like health centres, churches, schools etc. NCG approach intervention consciously improved beneficiaries understanding of safe MHM practices. Some MHM challenges persist such as lack of readily access to MHM materials due to non-availability and non-affordability, burden of managing menstrual pains and illnesses, and menstrual taboos. Yet, it appears the intervention is successful in bringing out the key unspoken MHM challenges in attempt to seek solutions although they are beyond the project scope and design. Most of the MHM challenges are systemic and/ or institutionalised rather than personal behaviour, even going beyond a single community. Menstrual stigma is comparatively very low and sometimes not recognized in some communities unlike the prevalence of menstrual taboos which are prominent across all communities (control and interventions). Menstrual taboos are directly linked to religious and superstitious beliefs while stigma could be triggered largely by a victim’s inappropriate behaviour and/ or poor personal hygiene practices. Championing local production and use of reusable menstrual pads from fabrics is an excellent intervention by the NCG project especially for women who could not afford commercial sanitary pads. It is recommended that MHM interventions should have enough scope and time to work directly with institutions and stakeholders especially those who have influence to facilitate long-term change processes like menstrual taboos with religio-cultural roots in local communities.

## Data Availability

All the relevant data supporting the findings have been shared directly in the paper. However, the raw data set could be shared under reasonable request.

## Acknowledgement

Authors appreciate the enormous support from World Vision US for funding this research work, and the National Team, especially the Area Program Offices in the Sekyere East District and Savelugu Municipal Assemblies particularly Rachel, Georgina, and Felix and our field assistants Timothy, Gideon, and Clement.

